# Self-determination theory in physiotherapy practice: A rapid review of randomized controlled trials and systematic reviews

**DOI:** 10.1101/2025.03.05.25323439

**Authors:** Jennifer O’Neil, Jacquie van Ierssel, Luc Pelletier

**Affiliations:** Faculty of Health Sciences, School of Rehabilitation Sciences, University of Ottawa, Ottawa, Canada; Bruyere Health Research Institute, Ottawa, Canada; Children’s Hospital of Eastern Ontario, Ottawa, Canada; Faculty of Social Sciences, School of Psychology, University of Ottawa, Ottawa, Canada

**Keywords:** Self-Determination Theory, Behaviour change, Service delivery, Physiotherapy

## Abstract

**Background:** The self-determination theory (SDT) is a theory on motivation proposing to support needs of autonomy, competence, and relatedness to improves autonomous motivation, which leads to adherence and compliance. Little is known about how SDT-driven physiotherapy interventions are implemented.

**Purpose:** The objectives of this rapid review were to identify the type of physiotherapy contexts in which SDT is being used and describe how SDT-based physiotherapy interventions are being measured.

**Methods:** The Cochrane Rapid Review Methods was followed to synthesize evidence from systematic reviews (SR) and randomized controlled trials (RCTs) on the use of SDT-related research in physiotherapy. We conducted a search on four databases between 1990 and September 17^th^, 2024. Two reviewers independently screened titles and abstracts, one reviewer completed the full-text screening while another screened all excluded full-text to ensure consensus. Findings were synthesized narratively following the review objectives.

**Results:** Of 184 identified SR or RCT, we included 8 RCTs (n=457) and 1 SR (n=712) targeting various health conditions for children and adults (i.e., cerebral palsy, adults with chronic condition). Physiotherapy interventions included strength and aerobic exercise, therapeutic modalities, yoga or tai chi, virtual therapy, coaching, and equine-assisted therapy. SDT techniques included communication training, autonomy supportive feedback, education and goal setting, provision of choices, and intrinsic motivation with the use of virtual reality, robotics, circus-themed games, music, and behaviour change strategies. A positive impact of SDT-driven physiotherapy interventions was seen for adherence, engagement, acceptance, physical activity, and intrinsic motivation.

**Conclusion:** Our rapid review suggests that SDT-driven physiotherapy is being used across a broad range of health conditions, using various physiotherapy and SDT principles derived from the theory.

## INTRODUCTION

Despite the importance of motivation in determining patient adherence to interventions and impact on treatment outcomes, little attention has been given to how motivational theory is being applied in physiotherapy practice. For physiotherapy interventions to be effective, a person must adhere to the proposed plan and complete the exercises suggested. Adherence to physiotherapy home exercise and management strategies throughout the duration of physiotherapy sessions is integral to short-term recovery, while long-term adherence can often reduce the incidence of recurrence or re-injury. Several intervention parameters such as duration, mode of delivery, cost, as well as behavioural factors including self-efficacy may influence adherence ^1^. Beyond the context of an intervention or behaviour of a client, a therapist’s behaviour may also have a significant impact on adherence. Precisely, interpersonal relationships, therapeutic relationship, shared-decision making are components that should be considered by a physiotherapist as they impact overall health outcome^2^. How a person behaves, in this case a physiotherapist, greatly influence motivation. Therapeutic relationships include person-centered care, interpersonal communication, and motivation strategies^2^ all components well reflected in the self-determination theory of motivation. According to the self-determination theory (SDT), interpersonal interactions are an integral part of the human experience that can directly influence the quality of the motivation of a targeted person, in this case a patient.

### Self-Determination Theory

SDT is a broad and comprehensive motivational theory developed by Deci and Ryan that has been widely used in psychology to inform interventions in the education, management, and health care sectors ^3,4^. According to SDT, motivation varies not only in function of quantity but also quality^5^. That is, motivation is not a simple dichotomous construct where a person is more or less motivated to engage in a behavior, motivation can take different forms that vary according to their level of autonomy or self-determination. SDT typically distinguishes between autonomous and controlled motivations^5,6^. Autonomous motivation refers to behavior that could be intrinsically motivated (a behavior done because it is pleasurable, that is an end in itself) and to behavior that could be extrinsically motivated (i.e., a mean to an end) but still self-determined. SDT refers to two types of self-determined extrinsic motivation, integrated regulation (where a behavior is further internalized by taking in consideration other dimensions of the self that are considered important) and identified regulation (where an internalized behavior is done by choice because it is considered important). Controlled motivation refers to behavior that are regulated by internal factors (like internal obligations, shame, guilt) that have been poorly internalized or external factors (like incentives, constraints, pressures from others). The more a behavior is autonomously regulated (or self-determined), as opposed to controlled (or non self-determined), it is likely to be integrated to a lifestyle and to lead to positive outcomes such as well-being^5,6^.

SDT proposes that for a person to be fully autonomous motivated and engaged in an activity, therapists should support the development of autonomous motivation from a self-determination stance by fulfilling basic psychological needs of: (1) autonomy (i.e., the need for individuals to act in line with their own interests and values), (2) competence (i.e., opportunities for increasingly challenging activities and developing greater levels of mastery), and (3) relatedness (i.e., needing a caring, supportive social network with strong interpersonal connections) ^4,7^. These three needs are said to be innate and universal across cultures. Need-supportive behaviours (i.e., supporting one’s autonomy, competence, and need for relatedness) promote need-supportive environments; while need-thwarting behaviours (autonomy thwarting/controlling, competence thwarting, and relatedness thwarting) lead to need-thwarting environments ^7^. According to SDT, if an individual’s context supports their basic psychological needs, their needs are more likely to be satisfied, which leads to autonomous motivation for a given activity. This allows them to engage in that activity out of interest or curiosity, or because it is in line with their goals and objectives ^7^. Alternatively, if an individual’s basic psychological needs are not met, they are more likely to be dissatisfied and engage in a behaviour for controlled reasons, such as to comply with or please someone else, or to avoid feeling guilt or shame. When individual engage in a behavior for controlled reasons, they are less likely to learn, to find the experience satisfying, and, in turn, to persist or maintain an activity. Since physiotherapy is often dependent on clients’ motivation and capacity for undergoing significant health behaviour change, a comprehensive review of how SDT improve patients’ active involvement with physiotherapy and facilitate health behavior change was necessary.

### SDT in physiotherapy

Autonomous motivation has been identified as an important contributor to physical activity adoption and intervention adherence^8,9^ and can enhance a patient’s active participation within medical encounters^10^. In terms of healthcare service delivery, needs-supporting relationships are as important when delivering services in-person as when using telehealth ^4,7^. Behaviours change techniques based on supporting basic psychological needs (autonomy, competence, relatedness) may present as goal setting, shared decision making, or occupational coaching in rehabilitation delivery. However, in rehabilitation-specific professions, the use of SDT is still in its infancy. In speech language pathology, a scoping review looking at aphasia and motivation identified a potential positive influence in clinical research for aphasia ^11^. In physiotherapy, the perception of a physiotherapist’s behaviour was shown to be important in the delivery of a telerehabilitation intervention ^12^. Autonomy, competence, and relatedness-supporting strategies, including, but not limited to, sharing of achievements, encouragement, and exchange of information, were systematically highlighted to be critical in peer support pain management rehabilitation groups ^13^. Although the influence of SDT has been indirectly studied in areas related to physical activity adherence, pain reduction, return to sport, and aphasia interventions, a gap still exist in understanding its specific use in physiotherapy intervention and how SDT is being delivered in that specific context ^9,14^.

According to the Canadian Physiotherapy Association, physiotherapy is “the assessment, diagnosis, and treatment of the human body, its diseases, disorders and conditions associated with physical function, and acute or chronic injury or pain. It includes the prevention of illness or injury and promotion and education on health and wellness with a focus on optimal movement and function” ^15^. Since prior physiotherapy research identified possible impact of components of self-determination theory implementation in a physiotherapy intervention^16^, we chose to document the precise use of this specific theory on the physiotherapy profession to facilitate reproducibility in a physiotherapy clinical context. In doing so, our aim is to examine the specific components of physiotherapy intervention that could be related to the satisfaction of the three basic needs proposed by SDT, that the behavior targeted by the intervention shows signs of internalization, and that this behavior is associated to the positive outcomes typically pursued by an effective physiotherapy intervention.

More specifically, the primary objective of this rapid review was to identify the physiotherapy contexts in which SDT is currently being used to determine whether using SDT principles within physiotherapy practice is effective at improving physiotherapy intervention adherence, facilitating education, and self-management. Our secondary objective was to describe the populations receiving SDT-based physiotherapy and how SDT-based physiotherapy interventions are being measured. Identifying this information will lead to the development of more effective SDT-driven physiotherapy interventions.

## METHODS

Rapid reviews improve accessibility and clarity of research evidence ^17,18^. Adhering to Cochrane methodology for rapid review ^19^, we examined how SDT approaches are being used within physiotherapy services to inform future program development. Based on high-quality evidence (i.e., RCTs and SRs), we wanted to identify the clinical application of theory to the population being targeted by this type of intervention and the methods of delivery in a physiotherapy context. We registered our protocol in the Open Science Framework repository before starting data extraction (https://doi.org/10.17605/OSF.IO/FP8JM). Specifics of the Cochrane methodology for rapid review are described in each methodology section.

### Search strategy

The search strategy was piloted in one database (i.e., MEDLINE) using Boolean logic “and/or” to combine specific MeSH terminology including study design (i.e., randomized controlled trial, systematic review), SDT (i.e., autonomy, determination, satisfaction, psychological needs, autonomous motivation), and physiotherapy (i.e., physical therapy, physiotherapy modalities, physical rehabilitation) (table in appendix). We subsequently expanded the search to include needs satisfaction in Cochrane Library (OVID), PsycInfo, and Embase databases between 1990 and January 2023. We updated the search in September 2024. We then hand searched the references of included full texts.

### Study selection

We screened citations using the review management software Covidence^TM.^ Two reviewers (JO, JvI) piloted a standardized title and abstract form for calibration before screening all articles by using the same 30 studies found in the initial MEDLINE search. As per Cochrane rapid review methodology, after duplicates were removed, two reviewers independently screened 50% of the titles and abstracts based on the following inclusion and exclusion criteria: systematic reviews and RCTs published in English and studies targeting physiotherapy services or exercise interventions based on SDT. Exclusion criteria will be incomplete studies, poster or protocols. In this review, the person who delivered the exercise program or physical activity was considered. Precisely, to be included, for an exercise program to be considered a physiotherapy intervention, the intervention needed to be delivered by a physiotherapist. Exercise or physical activity interventions were excluded when they were delivered by other health professionals or when the provider was not specified.

Once consensus was achieved, one reviewer (JO) completed the review of all titles and abstracts. Our inclusion criteria were SRs and RCTs published in English between 1990-2024, targeting physiotherapy services (i.e., evaluation, interventions, service delivery) or exercise interventions provided by a physiotherapist, and used SDT in the development of the interventions or assessments. Protocols and observational studies were excluded. Using the same process as above, we screened 10 full-text studies to pilot the full-text reviewing form. One reviewer (JO) completed the screening of all full texts while another reviewer (JvI) independently screened all excluded full text. Conflicts were resolved by discussion between reviewers until consensus was reached.

### Data extraction and synthesis

Data extraction was completed using a standardized form (appendix B) to extract how physiotherapy interventions and SDT techniques (i.e., communication, education, training program, behaviours, were delivered (i.e., in-person, telerehabilitation, hybrid), types of physiotherapy outcomes (i.e., adherence, effectiveness, health-equity) and SDT psychological needs(i.e., autonomy, competence, relatedness motivation, adherence, effectiveness, health-equity) being targeted, and the population and types of health conditions being investigated. The behavioural change technique classification of the data was informed by Teixeira et al. (2020)^20^. Data was extracted by one reviewer (initials here-anonymous) and reviewed by a second (initials here-anonymous). Two independent reviewers assessed Risk of Bias using the ROB2 tool ^21^ for RCTs and the AMSTAR2 tool ^22,23^. We followed the PRISMA reporting guidelines ^23^. Findings were analyzed narratively following the review objectives to identify similarities and differences between studies and synthesized descriptively in tables ^24^.

## RESULTS

### Included studies

Of 184 citations identified, we removed 58 duplicates, screened 126 titles and abstracts, reviewed 54 full-texts, and included 8 RCTs (n= 445 participants) and 1 SR (n=712 participants) (table 1). Interrater reliability for titles and abstracts selection was strong (k 0.88). The 9 studies retained in this review represented 1157 participants with an average age ranging from 3 to 77 years old. Only one systematic review targeted people under the age of 18 ^25^. The terminology for sex and gender was often interchanged, leaving no explicit reporting of gender in any of the studies. The four most recent RCTs ^26–29^ published between 2022 and 2024 reported more health equity indicators (i.e., socio-economic factors, employment, level of education, gender, race, and culture) compared to the four published earlier.

**Table 1.** Physiotherapy interventions, intervention delivery, and outcomes. Abbreviations: *HIV*, human immunodeficiency virus; *RCT*, randomized controlled trial; *SDT*, self-determination theory.

| Author, year, and sample size | Physiotherapy interventions | Intervention outcomes (i.e., adherence, engagement effectiveness, health-equity) | Mode of delivery |
| --- | --- | --- | --- |
| **Systematic Review (N=1) including 3 RCT, 6 Case-control, 6 non-comparative studies.** |  |  |  |
| **Meyens, de Mettelong, van der Spank et al. 2018.**<br>**Systematic review (n=712)** | Various physiotherapy interventions including virtual reality based therapy to train balance and strength (8 studies), Modified constraint-induced therapy (1 study), supported treadmill training (1 study), Robotic (1 study), circus-camp (1 study) for children with developmental disorders. Parameters ranged from 1 day 12 minutes session to 20 weeks. | **Compliance:** VR is an innovative or fun aspect to increase the participants' exercise time or compliance (12 studies).<br>**Physical Activity:** exergames did not provide as vigorous physical activity training as the physical fitness exercises (1 study).<br>**Engagement:** VR intervention was enjoyable and motivating for children with CP | Various modes of delivery including in-person, virtual reality, robot-assisted. |
| **Randomized controlled trials (RCTs) (n=8)** |  |  |  |
| **Murray et al. 2015**<br>**Randomized controlled trials (n=48)** | Communication skill training for physiotherapist working with adults with chronic back pain | No outcome on patients | In-person physiotherapy delivery in outpatient clinics. |
| **Shah, Majeed, Yoruk, et al. 2016**<br>**Randomized controlled trials (n=67)** | Individually tailored 12-weeks community-based physical activity program that included walking at moderate intensity and 10 tailored, moderate to intense resistance strength training exercises for upper and lower body ranging from 1-4 set of 8-15 repetitions with exercise training bands. Adults with HIV-infected at risk of functional decline. | **Physical activity:** Significant improvements in physical function and activity within the treatment group.<br>**Depression:** Significant improvement in emotional well-being and role limitations due to physical health in the treatment group compared to the control group. | Recommended by an experienced physical therapist |
| **Levac et al. 2017**<br>**Randomized controlled trials (n=40)** | No physiotherapy intervention for patient provided | **Engagement:** Participants in the autonomy-supportive group demonstrated better retention (i.e. shorter movement time) on sequence. There were no significant differences between groups in overall or subscale user engagement scale. | Virtual reality delivered in a research laboratory. |
| **Aunger et al. 2020**<br>**Randomized controlled trials (n=35)** | Physiotherapy intervention for older adults waiting for hip or knee replacement surgery | **Adherence:** 88% completion rate<br>**Physical activity:** Eight (42.1%) achieved or exceeded their step targets, usual care (4907 mean steps per day) vs intervention group (5170 mean steps per day). | In-person physiotherapy delivery. |
| **Wallbank et al. 2022**<br>**Randomized controlled trials (n= 62)** | The Active Women over 50 Online Program offered over 3-months in the community. | **Acceptability:** 83% participants would recommend participation to another person | Online program offered in the community. |
| **Quinn et al. 2022**<br>**Randomized controlled trials (n=57)** | The PACE-HD is a physical activity program that includes 18 sessions of 30-minutes of moderate-vigorous exercise, 3–5 times per week for adults living with Huntington’s disease. | **Adherence:** 80.5% adherence<br>**Physical activity:** Differences walking endurance and self-reported PA at 12 months favoring the intervention group | In-person or videoconference by a physiotherapist. |
| **Chair et al. 2024**<br>**Randomized controlled trials (n =130)** | An 8-week hybrid program including 16 supervised center-based exercise sessions (2 hours/session, 2 sessions/week) of prescribed exercises and home-based self-paced program for adults living with coronary heart disease. | **Exercise capacity:** The intervention group demonstrated statistically significantly greater improvements in exercise capacity compared to control group. | Hybrid program |
| **Nunez-Gaunard et al. 2024**<br>**Randomized controlled trials (n = 18)** | A 12-week intervention including aerobic and strengthening exercises once a week with progressions during week 3 to 10 for adults between 18 and 32 years of age with a diagnosis of autism spectrum disorder. | **Adherence:** Attendance to weekly telehealth sessions ranged from 70 to 100%, with a mean of 93%. Wearable adherence ranged from 20 to 100%, with a mean of 82%.<br>**Physical Activity:** Significant effect from the intervention for physical activity pre-post-test and based on steps/day | Telehealth-synchronous program |

### Risk of Bias

Overall, two of the RCTs were rated as low ROB while 6 RCTs were rated as having some concerns (figure 2). Concerns were mostly related to the randomization process and missing data. The SR by was assessed as high quality using the AMSTAR 2 tool ^22^, however, the lack of protocol registration could introduce bias.

**Figure 1.**
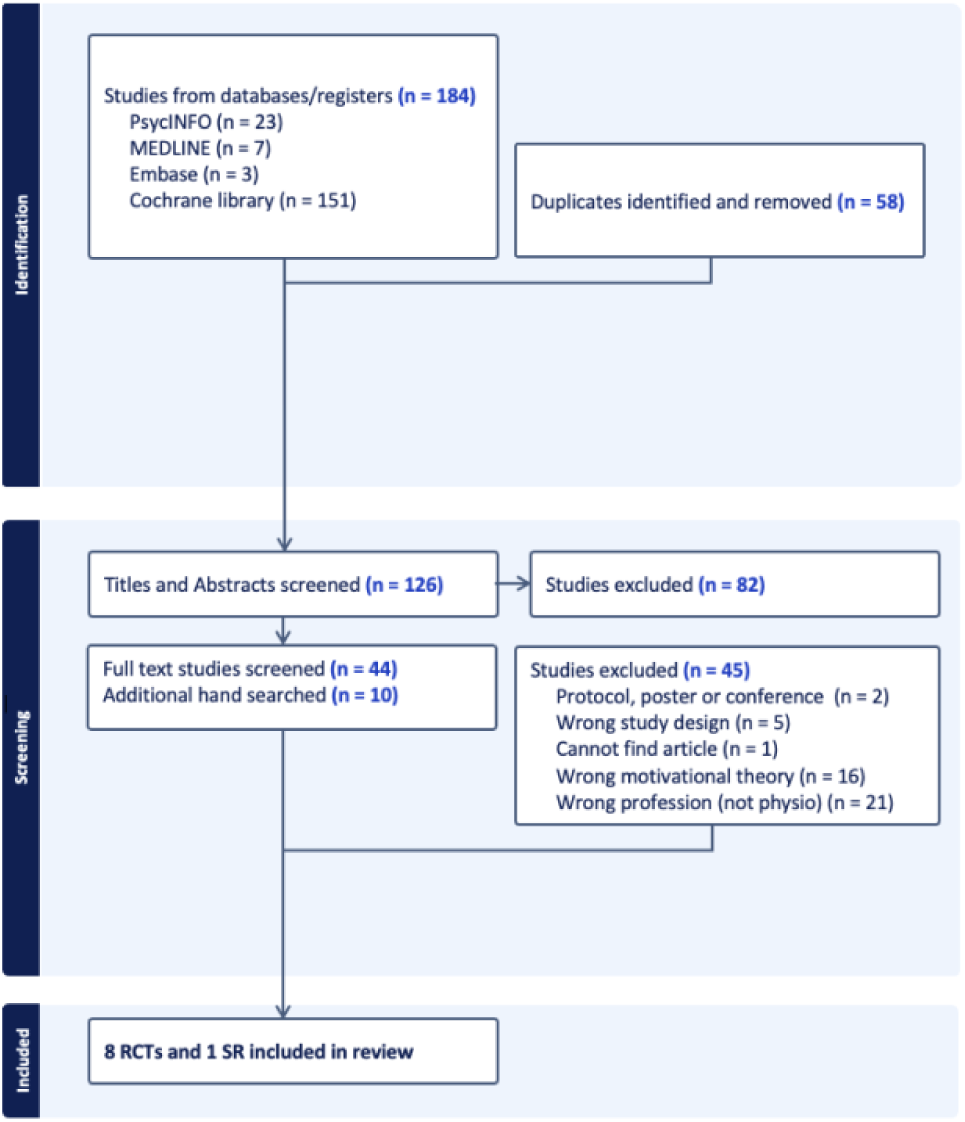
Prisma Chart illustrating the identification, screening, and inclusion process^23^.

**Figure 2.**
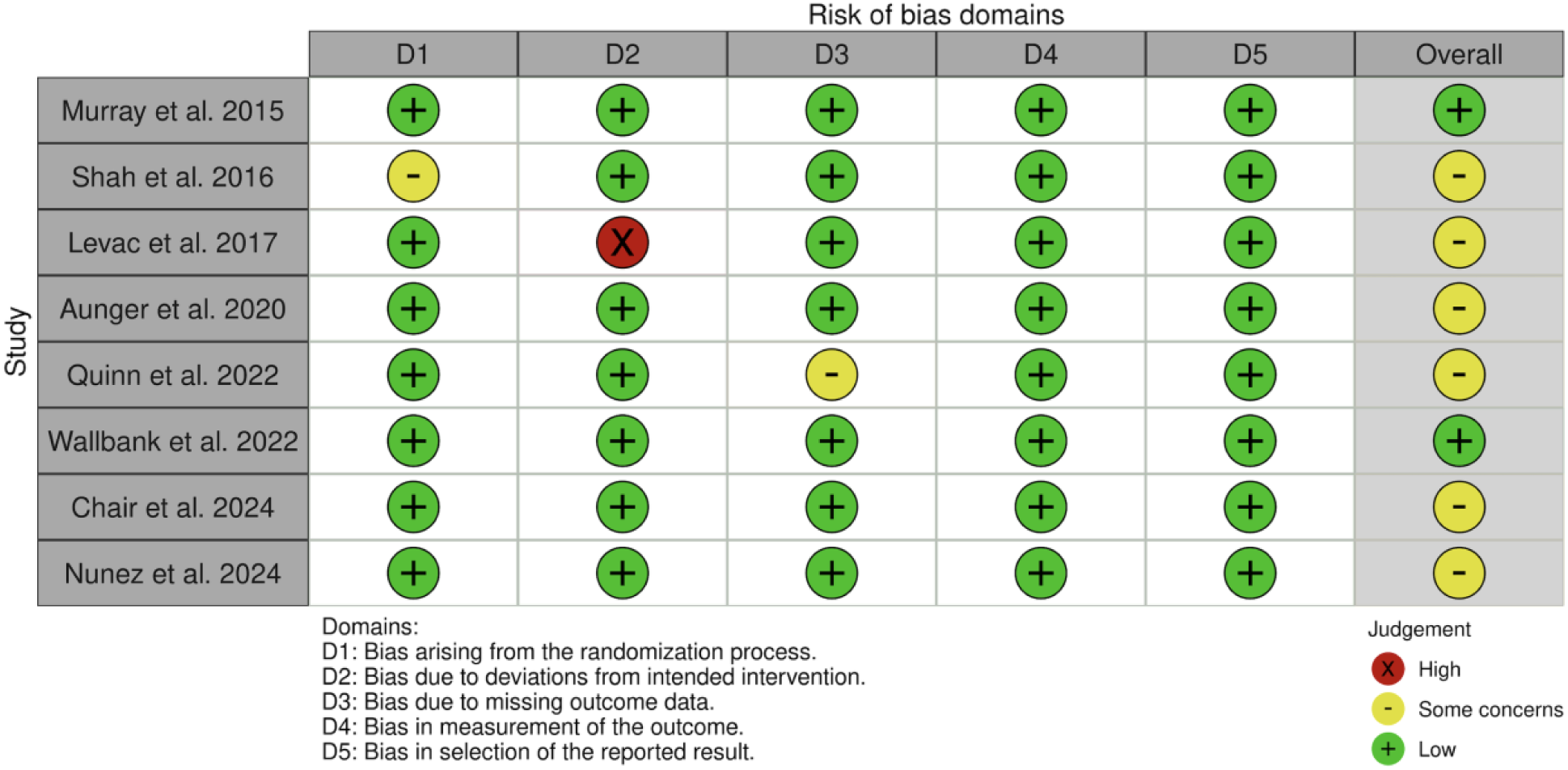
Reporting on Risk of Bias (ROB) of the RCTs assessed by the ROB2 tool^30^.

### Physiotherapy and SDT interventions

Patient populations included adults living with specific orthopedic conditions (including chronic low back pain, those requiring hip and knee replacements, and sedentary women) and adults and children with specific neurological conditions (including HIV, Huntington disease, and children with motor development conditions such as cerebral palsy, brachial plexus injuries, and gait development impairments, and autism spectrum disorder). One RCT targeted adults with coronary heart disorder and one was conducted with healthy students (table 1).

Several types of physiotherapy interventions for children and adults were included such as traditional strength and aerobic exercise programs ^26–28,31–34^, virtual reality therapy ^25,35^, and equine-assisted therapy^25^. Diverse settings included outpatient hospitals ^32,34^, hybrid interventions ^26^, home and community ^25–29,34,36^ and research laboratories ^35^ (table 1).

The SR by Meyens et al. focused on delivery of different motivational techniques including the use of virtual reality, robotics, circus-themed games in a goal-based approach to improve intrinsic motivation in children with developmental disorders^25^. Five out of eight RCTs used multifactorial motivational strategies including goal setting, education on physical activity, exploration of barriers and facilitators, problem solving, adherence, social interaction, and self-monitoring ^26–29,34^. Another RCT^35^ conducted with healthy students in laboratory environments used virtual reality to compare autonomy-supportive and autonomy-controlling tasks. In contrast, Murray et al. (2015)^32^ used hybrid behavioural change techniques and methods of delivery, including video recording, vignettes, active role-playing, and group discussions to enhance the physiotherapist’s abilities to support their patient’s needs. Novel combinations of in-person, group videoconference, and phone applications ^27^, the benefit of music and phone calls ^26^, and phone interviewing and text-messages ^29^ were also documented. While other RCTs relied on a combination of simple methods, including in-person and phone calls ^28,33^, Aunger et al. (2020) only included in-home, in-person motivational strategies ^34^.

### Outcomes and outcome measures

RCTs measured outcomes related to physical activity and function, behaviour changes, mental health outcomes, quality of life, and a combination of these (table 3). Specific to physical activity level, seven RCTs ^26–29,33–35^ used a variety of outcome measures, including self-reported questionnaires, clinical assessment, and objective kinematic measures. Five RCTs ^26–29,34^ used wearable technology to capture step count and overall daily physical activity in real-world context. Other physical outcomes and outcome measures included strength measured by dynamometer ^33^, function measured by daily repetitions of sit-to-stand ^34^, and overall disability ^29^.

**Table 2.** Motivation Behaviour Change Techniques (MBCT), SDT intervention and delivery and outcomes. Abbreviations: MBCT - Motivation Behaviour Change Techniques, SDT – Self-determination theory. Note: The number beside MBCT refers to the Classification of Motivation and Behavior Change Techniques label/taxonomy.

| **Author, year, and sample size** | **Motivation Behaviour Change Techniques (MBCT) and SDT interventions** | **Outcomes** | **Mode of delivery** |
| --- | --- | --- | --- |
| **Meyens, de Mettelong, van der Spank et al. 2018.**<br>**Systematic Review (n=712)** | 12 out of 15 studies included fun aspects targeting intrinsic motivation. This included VR games for motivation for motor rehabilitation (5 studies), | **Intrinsic motivation:**<br>The authors found that exergames initially had a high intrinsic motivational power with high situational interest that attracted the children (lasting effects of Challenge, Exploration and Instant Enjoyment). This indicates that a diversion from the exercise (movie) provides a similarly increased motivation to “go through” with the rehabilitation as being immersed in the VR game to increase the intrinsic motivation actively participate in the rehabilitation. | Virtual reality as mode of delivery. |
| **Murray et al. 2015** | **1. Competence-support techniques**<br>**MBCT 18. Offer constructive, clear, and** | **Needs-supportive behaviours:** A large difference in needs-support scores between physiotherapist in | Hybrid delivery using videorecording, vignettes, active role-playing, |
| **Randomized controlled trials (n=48)** | **relevant feedback.**<br>Eight-hour communication skill training including SDT concepts designed for physiotherapists. | intervention group physiotherapists rated as significantly more supportive than control arm physiotherapists. | and group discussion. |
| **Shah, Majeed, Yoruk, et al. 2016**<br>**Randomized controlled trials (n=67)** | **1. Autonomy supportive interventions**<br>**MBCT1. Elicit perspectives on condition or behavior, MBCT 4. Explore life aspirations and values**<br>Six patient-centered counseling sessions: 60 minute in-person counseling session on individual's interests, values, and behaviours sessions for two weeks.<br>**MBCT 6. Provide choice**<br>Participants also took an active role in the customization of their own PA program.<br>**2. Competence-support techniques**<br>**MBCT 15. Address obstacles for change**<br>15-30 minutes follow-up telephone counseling sessions every two weeks to provide positive feedback, re-evaluate and modify goals. | **Intrinsic motivation:**<br>No clear evidence of treatment effects on, perceived competence, or intrinsic regulation, however, participants who improved in intrinsic motivation also improved in gait speed. | 60 minute in-person counseling session and telephone counseling sessions |
| **Levac et al. 2017**<br>**Randomized controlled trials (n=40)** | **1. Autonomy supportive interventions**<br>**MBCT2. Prompt identification of sources of pressure for behavior change and MBCT 7. Encourage the person to experiment and self-initiate the behavior**<br>Virtual reality system in the Rehabilitation Games and | **Intrinsic motivation:**<br>There was a significant difference between groups in overall IMI score with the autonomy supportive group performing higher on overall motivation. | In a research laboratory |
|  | Virtual Reality intervention for physiotherapists using the force plate and a 180-degree flat projection screens with external speakers to convey auditory feedback. Participants were randomized into autonomy-supportive (OPTIMAL) or autonomy-controlling conditions and to sequence practice order using a random number generator (Locus of control). |  |  |
| **Aunger et al. 2020**<br>**Randomized controlled trials (n=35)** | **1. Competence-support techniques**<br>**MBCT 17. Assist in setting optimal challenge, MBCT 19. Help develop a clear and concrete plan of action, and MBCT 20. Promote self-monitoring.**<br>Formulation of action and behavioural goals, illumination of discrepancies, problem-solving, reframing, and self-monitoring.<br>**2. Relatedness-support techniques**<br>**MBCT 9. Encourage asking of questions**<br>Behaviour changes interventions including motivational interviewing, emotional support, health and physical education. | No specific behaviour change outcomes reported | Delivered in 2 visits in the person's home. |
| **Wallbank et al. 2022**<br>**Randomized controlled trials (n= 62)** | **1. Relatedness-support techniques**<br>**MBCT 13. Providing opportunities for ongoing support**<br>Health coaching messages including benefits of physical | **Acceptability:**<br>87% participants chose to receive motivational messages, 79% opened email messages sent, 31% of the embedded links were clicked on, and 88% of participants | Emails and SMS messages (as per patient preference) delivered via telephone by a tertiary-qualified physiotherapist |
|  | activity, social modeling, ways to address barriers to activity, and help with action planning. | indicated the telephone health coaching session supported their PA. | trained in motivational interviewing. |
| **Quinn et al. 2022**<br>**Randomized controlled trials (n= 57)** | **1. Autonomy supportive interventions**<br>**MBCT 7. Encourage the person to experiment and self-initiate the behavior**<br>Eighteen physical activity coaching sessions including physical activity coaching, and engagement strategies.<br>**2. Competence-support techniques**<br>**MBCT 20. Promote self-monitoring**<br>Goals monitoring. | **Acceptability:**<br>Participants reported a very high level of satisfaction with 20/20 (100%) agreeing or strongly agreeing that they were satisfied by and enjoyed the therapist-led sessions. | Delivered on the phone by a site coordinators or licensed physical therapists. |
| **Chair et al. 2024**<br>**Randomized controlled trials (n =130)** | **Autonomy supportive interventions**<br>**MBCT 7. Encourage the person to experiment and self-initiate the behavior**<br>Patients completed 135 moderate-intensity tempo songs in addition to a music-paced physical activity developed by a musician with a bachelor's degree in music.<br>**MBCT 4. Explore life aspirations and values**<br>5 biweekly telephone-based sessions including discussion around adherence, internal conflict related to physical activity, perceived benefits and barriers. | **Self-efficacy:** The intervention group had significantly improved exercise self-efficacy compared to control group. | In-person and telephone-based |
| **Nunez-Gaunard et al. 2024**<br>**Randomized controlled trials (n = 18)** | **1. Competence-support techniques**<br>**MBCT 20. Promote self-monitoring**<br>Exercises progress monitoring, goal reflection around physical activity education.<br>**2. Relatedness-support techniques**<br>**MBCT 14. Prompt identification and seek available social support**<br>Weekly online discussions during 10 weeks with fitness support Pods, including success celebrations, problem-solving, and social relatedness using the Fitbit app. | Relatedness, competence, and autonomy supportive | Weekly individual and group online discussion for 10 weeks. |

**Table 3.** Outcomes measures used in physical or behaviour change domains. Abbreviations: *EuroQol EQ-5D-5L*, European Quality of Life – 5 Dimensions, 5 Levels; *ISWT*, Incremental Shuttle Walk Test; *PA*, physical activity; *WHO*, World Health Organization.

| References, date | Physical outcomes and outcome measures | Behaviour changes, mental health outcomes, and outcome measures | Intervention feasibility and fidelity outcomes and outcome measures |
| --- | --- | --- | --- |
| **Systematic Review** |  |  |  |
| **Meyens et al. 2018** | Not applicable | **Motivation and engagement:** Pediatric Volitional Questionnaire, Pediatric Motivational Scale, | **Satisfaction:** Likert scale assessing fun in rehabilitation |
|  |  | Children's Assessment of Participation and Enjoyment, Situational Interest Scale-Elementary school, Family Participation |  |
| **Randomized Controlled Trials** |  |  |  |
| **Murray et al. 2015** | Not applicable | **Needs-supportive behaviours:** Health Care Climate Questionnaire | Not applicable |
| **Shah et al. 2016** | **Physical function and physical activity:** Physical Performance Test, a MicroFET 2; time needed to complete an obstacle course; six-minute walk test; gait speed; 12-item Physical Activities Scale for the Elderly questionnaire; Paffenburger Physical Activity Index<br>**Physical function (strength):** hand-held dynamometer | **Motivation and engagement:** 3-item Locus of Causality for PA scale; Behavioral Regulation in Exercise Questionnaire-2; Perceived Competence scale is a 4-item scale<br>**Mental health:** the Beck Depression Inventory-II,<br>**Quality of life:** the Short-Form Health Survey-36 | Not applicable |
| **Levac et al. 2017** | **Physical function:** Acquisition and Retention time | **Motivation and engagement:** Intrinsic Motivation Inventory; User Engagement Scale | Not applicable |
| **Aunger et al. 2020** | **Physical activity levels:** ActivPal3 to measure sedentary time, steps per day, measure of Older Adults Sedentary Time, Short Physical Performance Battery<br>**Physical function (strength):** daily sit-to-stand | **Motivation and needs supportive behaviours:** Basic Psychology Needs Scale | **Adherence and compliance:** adherence to action plan in booklet<br>**Intervention fidelity:** rating of delivery skills (support of basic psychological needs, motivational interview, problem solving, progress monitoring, setback management) |
| **Wallbank et al. 2022** | **Physical activity levels:** | **Motivation and Exercise efficacy:** | **Acceptability:** whether participants |
|  | average number of daily steps measured by ActiGraph GT3X+ accelerometer; self-reported physical activity, measured by the International Physical Activity Questionnaire; proportion of participants meeting at least the lower limits of the WHO physical activity guidelines; 150 min/week of moderate-intensity physical activity or 75 min/week of vigorous-intensity physical activity or an equivalent combination of both, measured by ActiGraph GT3X+ accelerometer<br>**Physical function:** function component of the Late Life Function and Disability Instrument. | Exercise perceptions, measured by the Exercise Benefits and Barriers Scale<br>**Mental health and mood:** the Positive and Negative Affect Schedule<br>**Health-related quality of life:** EuroQol EQ-5D-5L survey | would recommend study participation to another person. |
| Quinn et al. 2022 | **Physical activity levels:** wearable activity monitors (Fitbit Charge 2) to monitor PA (e.g. step counts, activity intensity and frequency). | **Mental health and behaviour:** Short Problem Behavior Assessment, Hospital Anxiety and Depression Scale combined with the Snaith Irritability Scale<br>**Health-related quality of life:** Short-Form Health Survey (SF-12) | **Adherence:** % of sessions attended<br>**Intervention fidelity:** therapist self-report checklists completed after each session and therapist self-assessment rating scale; average number of days participants had Fitbit devices and % of days with valid wear time (defined as $\geq 10$ h)<br>**Safety:** calculating the number of adverse events and number of falls in intervention and control groups |
| **Chair et al. 2024** | **Physical Activity Levels:** the 10-m ISWT; heart rate using a sensor (Polar H7, Polar Electro GmbH, Germany); blood pressure and heart rate before and after the test; perceived exertion at the beginning and end of the test; accelerometer for 7 days; the validated International Physical Activity Questionnaire-Short Form | **Exercise self-efficacy and self-determination:** the validated Chinese version of the Cardiac Exercise Self-Efficacy Instrument; Behavioural Regulation in Exercise Questionnaire. | Not applicable |
| **Nunez-Gaunard et al. 2024** | **Physical activity levels:** Godin-Shephard Leisure-Time Physical Activity Questionnaire, Fitbit Inspire (Fitbit® Inc., San Francisco, California, USA), for 10-weeks.<br>**Adiposity Anthropometric assessments:** body mass index; waist to height ratio via telehealth | **Exercise self-determination:** Behavioral Regulation in Scale; relative autonomy index | **Adherence:** attendance to session<br>**Intervention fidelity:** wearable adherence<br>**Safety:** adverse events related to music-paced physical activity |

Outcomes related to needs-supportive behaviour changes and mental health were mostly measured using questionnaires and validated scales. Motivation and engagement were measured in the SR (Meyns et al., 2018) and six RCTs ^26,27,29,33–35^. The most frequently used measure for motivation and engagement was the Behavioral Regulation in Exercise Questionnaire (3 out 9 studies). Changes in mood and mental health were measured in three RCTs ^28,29,33^ with the Beck Depression Inventory, and the Positive and Negative Affect Schedule. Finally, quality of life was measured in three RCTs ^28,29,33^ using three different measures, the Short-Form Health Survey-36, the Short-Form Health Survey-12, and the EuroQuol EQ-5D-5L survey.

Adherence or attendance was measured differently across studies, including reporting of adherence to action plans in a booklet (comprised of six goals and three environmental modifications) ^34^ and percentage of sessions attended ^27,28^. Intervention fidelity was measured in 3 RCTs by rating the delivery of skills ^34^, self-reporting checklist from physiotherapists ^28^, and use of wearable sensors ^27,28^. Two RCTs measured safety, and one RCT measured satisfaction and acceptability.

### Efficacy of physiotherapy services combined with SDT interventions

One RCT reported a significant increase in physiotherapists’ needs-supportive behaviours with SDT interventions measured by the Health Care Climate Questionnaire (intervention arm [mean ±SD] = 4.6±0.85; control arm [mean ±SD] = 2.78 ±0.72)^38^. In terms of intervention feasibility and acceptability, one RCT reported that combining in-person physiotherapy services with motivational interviewing, emotional support, education, and problem-solving is feasible and may have an impact on sedentariness in older adults awaiting hip and knee arthroplasty ^34^. Another RCT demonstrated that combining online physiotherapy program with health coaching emails and text message was feasible and an acceptable for women over 50 years old ^29^. A third RCT showed that combining moderate-vigorous exercise 3–5 times per week with 18 physical activity coaching sessions may have a positive impact on walking endurance and self-reported physical activity in people living with Huntington’s disease ^28^. Findings also suggest that combined physiotherapy interventions with SDT-based interventions could significantly improve physical function such as gait speed in people with HIV ^33^, exercise capacity and exercise efficacy in people with coronary heart disease ^26^, and self-reported physical activity in adults with autism spectrum disorder ^27^.

## DISCUSSION

In this rapid review, we describe the use of SDT techniques combined with physiotherapy interventions, how they are measured, and their reported outcomes based on high quality evidence. Our findings also provide some insight on how physiotherapy interventions combined with SDT techniques are currently being delivered (e.g., aerobic exercise, virtual reality, problem-solving tools, reframing), how are these interventions currently being assessed (e.g., physical activity levels, motivation, mood and behaviour), and for whom they have been designed. Overall, our findings suggest that SDT techniques combined directly with specific physiotherapy interventions is feasible for people with a variety of health conditions, including children with neurodevelopmental disorders such as cerebral palsy, adults with certain neurological conditions including HIV and Huntington’s disease, adults waiting hip or knee replacement or who are sedentary, and adults with cardiovascular conditions. Studies on the impact of physiotherapy and SDT interventions developed for adults and older adults with neurological conditions such as stroke, traumatic brain injury, spinal cord injury, and neuroinflammatory conditions are still lacking^39,40^. Since research shows that personalization and tailoring of physiotherapy interventions based on the SDT framework positively impact motivation in people who have experienced a stroke ^41^, however the current literature does not report enough details on the types of tailoring or motivational strategies to enable clinical recommendations or replication^42^. It is necessary to understand the specific motivation and behaviour change techniques used within specific contexts and with specific populations to tailor care.

This review also showed that supporting competence of physiotherapist using SDT techniques may indirectly influence communication skills of physiotherapists, which may improve therapeutic relationships between clinicians and patients^12^. Furthermore, results from cohort studies included in Meyns et al. suggest that the use of virtual reality as well as meaningful partnerships may improve active engagement within a physiotherapy and effectiveness of the intervention ^43–46^. The impacts of combining SDT specific behaviour change techniques within a physiotherapy framework may have an impact on both, clinicians and patients. Consistent with findings from a meta-analysis by McGrance et al., motivational SDT interventions included in this rapid intervention primarily improved adherence, exercise behaviour, and levels of physical activity ^47^. From a motivational and behaviour change perspective, results from this rapid review also showed positive outcomes related to clinicians’ needs supportive behaviours as well as patients’ self-efficacy and acceptance. The challenges now lie with knowledge of such complex interventions and how to implement them clinically. Approaches such as co-creation of interventions or shared-decision activities might be beneficial^10^. Training of future physiotherapist on the implementation of behaviour change within a clinical context is critical.

Furthermore, our results suggest that is feasible for physiotherapy interventions combined with behaviour changes and SDT techniques to be delivered in various modes. It demonstrates the potential of alternative physiotherapy delivery options (i.e., in-person, telerehabilitation, robot-assisted, hybrid) and SDT techniques delivery options (e.g., in-person, telerehabilitation, virtual reality, phone sessions, text-messages). When comparing in-person and telerehabilitation in physiotherapy, emerging evidence shows similar results for satisfaction, adherence, and effectiveness for certain conditions ^48–50^. Our findings showcase the acceptability and effectiveness of diverse physiotherapy care delivery. Similarly, the delivery of interventions, both physiotherapy and SDT, was reported to be feasible and accepted in various settings (hospital, virtual reality, research laboratory, vignettes). Further high-quality research is needed to explore how physiotherapy and behaviour change techniques can be integrated in complex interventions, to identify if the method of delivery has an impact on outcome, and how the method of delivery may influence the design of complex telerehabilitation interventions.

Interesting findings around outcome measures is highlighted. Our rapid review demonstrated a lack of standardized outcome measures related to autonomous motivation, communication and interpersonal behaviours and their validation for physiotherapy interventions. This is an important clinical and research gap since autonomous motivation has been demonstrated to have an impact on health behaviour^51^. Assessment tools objectively measuring the impact of SDT combined with physiotherapy interventions may be necessary to demonstrate the effectiveness of SDT-driven physiotherapy. Reliance of self-report may create limitation in the assessment of effectiveness of SDT-driven physiotherapy. Rouse et al. provide an example of a validated tool measuring the interpersonal support in physical activity consultation^52^. Others have proposed self-determination specific questionnaires such as the Health Care Climate Questionnaire, the Perceived Competence Scale, and Treatment Self-Regulation Questionnaire^53^. Without access to or use of validated tools designed to measure if and how a motivation-based intervention may influence physiotherapy interventions, the inclusion of SDT interventions within physiotherapy may be limited. This review could influence the development or clinical use of new self-determination-based outcome measures that are simple to use in all geographical context and languages.

Finally, demographic findings demonstrate gaps in explicit reporting of health equity indicators. Reporting of health equity indicators could not only improve transparency in reporting but provide critical information on the context in which the person is living ^54^ for whom a combined physiotherapy and SDT intervention would be beneficial. Results from our updated search demonstrates a positive trend in the reporting of key socio-demographic indicators. Future work should document in detail how health equity indicators are considered and how SDT interventions and supporting basic psychological needs could influence the effectiveness of an intervention from a sex and gender-based analysis, a cost-effectiveness analysis, or an accessibility analysis.

### Practical Implications

1. Supporting basic psychological needs combined with physiotherapy interventions may have a positive impact on a clinician’s therapeutic approach as well as certain patient health outcomes, adherence to intervention, and the development of effective therapeutic relationships. Behaviour change techniques should be systematically included as part of the physiotherapy curriculum as well as a continuing education topic for current physiotherapists, focusing on hands-on implementation rather than theoretical ideas.
2. Combining physiotherapy interventions and SDT techniques is feasible and acceptable for various health conditions within various health care settings. It is therefore important to document how these interventions have been implemented and integrated into new interventions or clinical practice to facilitate implementation.
3. Combining physiotherapy interventions (i.e.., moderate to high intensity resistance training, aerobic training, constraint–induced movement therapy) and SDT techniques (virtual reality, person-centered coaching sessions, music) can improve adherence to intervention, physical activity levels, and intrinsic motivation. Physiotherapists should strive to combine these interventions tools to improve self-efficacy and consequently efficacy of interventions.

### Limitations

We acknowledge that the exclusion of other motivational theories may limit the generalization of our results. Future reviews may want to include other motivational theories, compare their impact, and synthesize their differences. It is critical to consider some limitations encountered during the screening and selection process of the studies. Unfortunately, the type of provider delivering the physical activity, physiotherapy of exercise program was often not reported. Since “physiotherapy or physiotherapist” was a specific inclusion criterion, many studies discussing ‘health or rehabilitation’ interventions were excluded. There is a need to report who is providing the intervention in studies to enable replication as well as personalize training around SDT for each health profession. The SDT interventions and behavioural change techniques presented in this review were not always reported based on the target behaviour change (competence, autonomy, relatedness support). Similar to documenting the details of physiotherapy interventions, precise reporting using validated measures of autonomous motivation or precise reporting of how the basic psychological needs of competence, autonomy, and relatedness may be supported or thwarted in physiotherapy, could impact clinical practice and effectiveness of physiotherapy interventions ^3^. Furthermore, due to the lack of consistency in outcome measures, a meta-analysis of results was not possible for this rapid review. It is therefore essential to consider using consistent validated measures, such as validated measures of physiotherapist’s needs-supportive and needs-thwarting interpersonal behaviours, and autonomous and controlled motivation for engaging in physiotherapy, when conducting effectiveness trials to allow for the development of robust clinical guidelines on SDT driven physiotherapy interventions. To mitigate this limitation, we narratively synthesized the results from each included study which allowed us to provide preliminary information on feasibility and effectiveness.

## CONCLUSION

With a growing push towards the use of person-centered approaches, goal setting, and shared decision making in physiotherapy, results from this rapid review demonstrate how SDT-designed physiotherapy intervention can be clinically implemented by identifying various types of SDT and physiotherapy interventions across various contexts. This review also identified key SDT interventions such as problem solving, problem monitoring, and goal setting which may, when combined with evidence-based physiotherapy interventions, improve the effectiveness of interventions. However, given the paucity of evidence of SDT interventions within physiotherapy, further research around implementation and delivery is needed before robust conclusions can be drawn as to the effectiveness on patient outcomes. This information may facilitate clinical implementation of SDT within physiotherapy, provide guidelines for the initiation and maintenance of health behaviours and specific adherence-interventions, as well as inform future training of physiotherapists.

## Data Availability

All data produced in the present study are available upon reasonable request to the authors

## Acknowledgements and Conflict of interest

This research did not receive any specific grant from funding agencies in the public, commercial, or not-for-profit sectors. The authors declare that they have no financial conflict of interest with the content of this article.

## Acknowledgements

NIL

## Appendix A

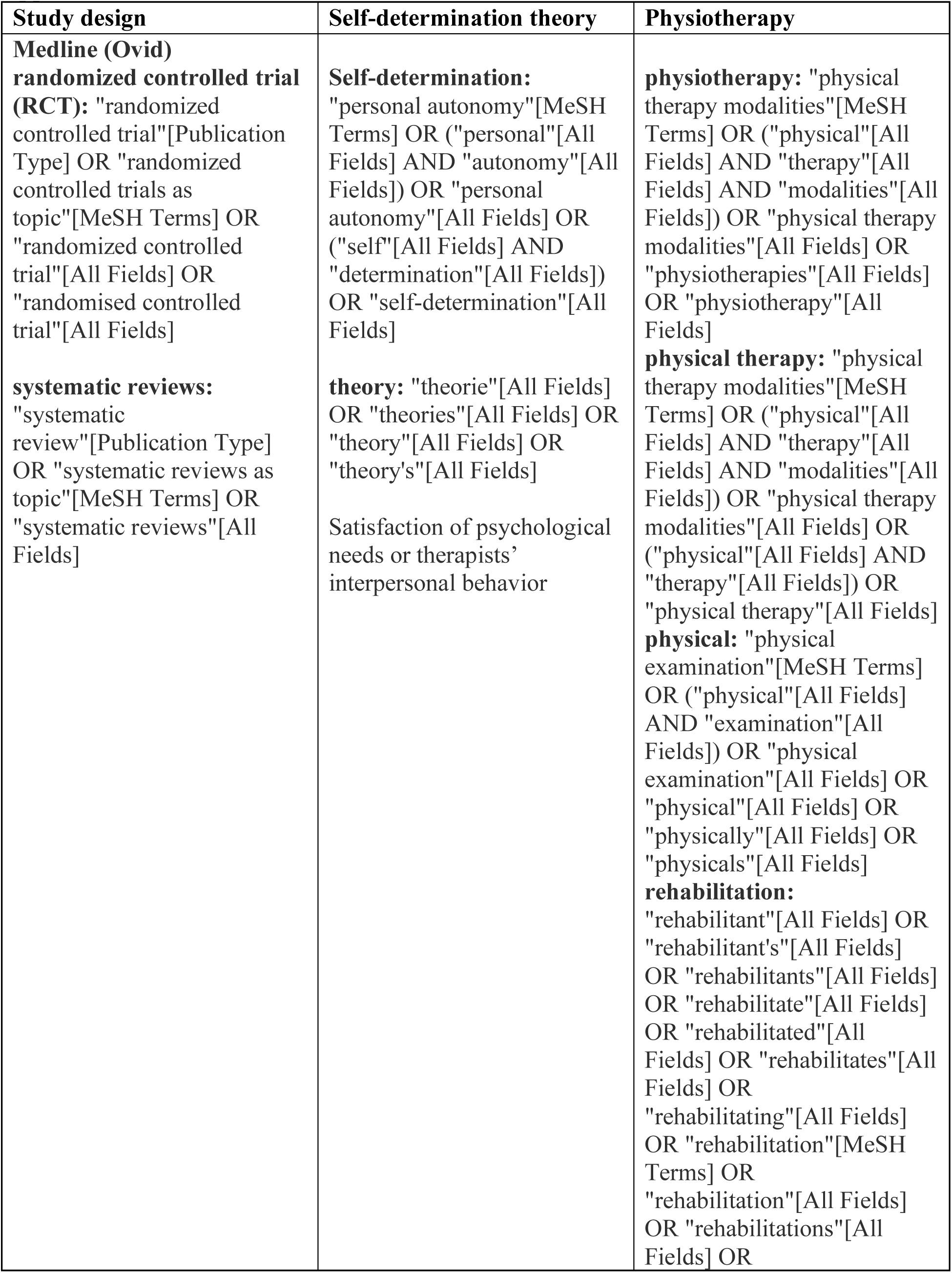
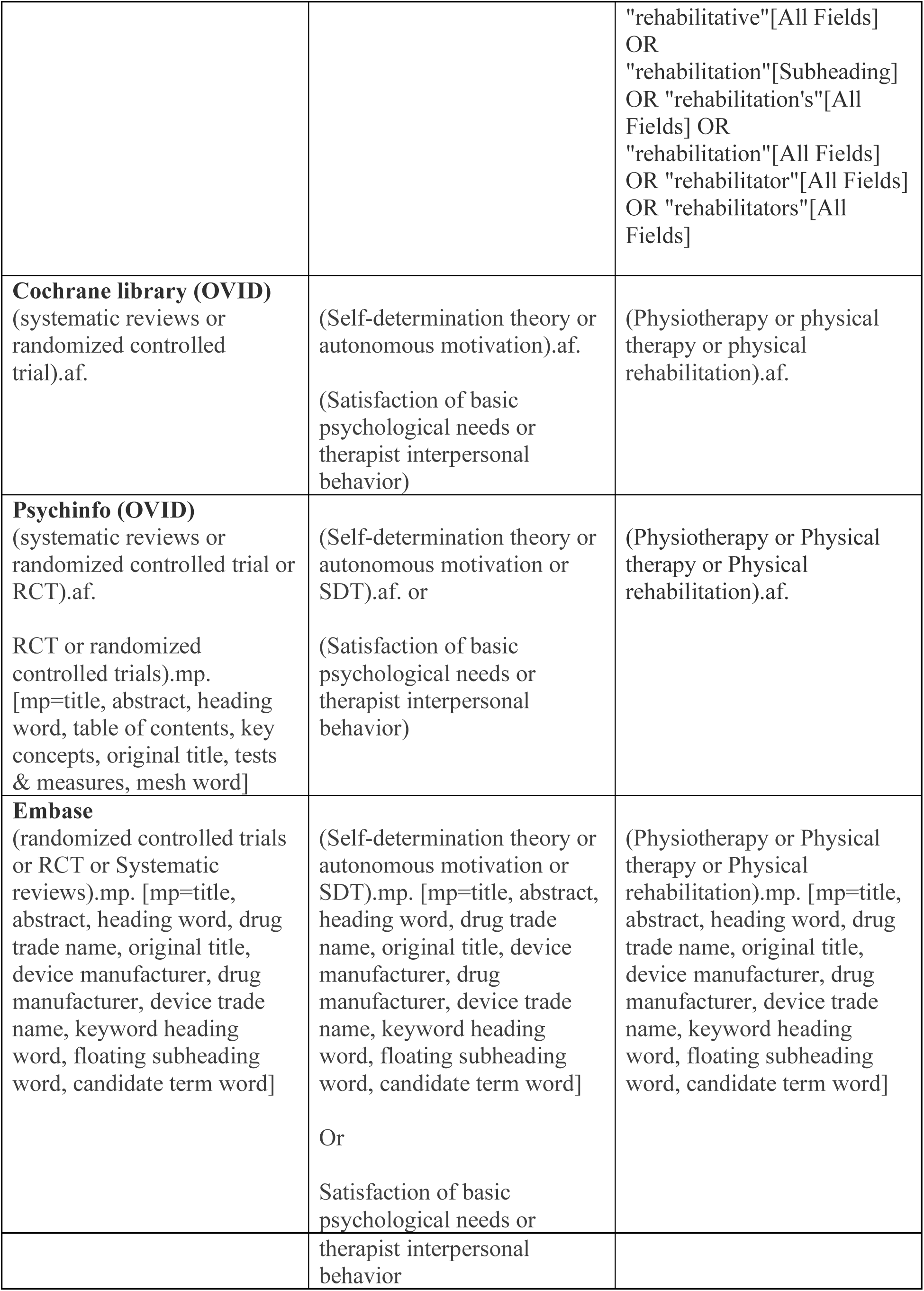

## Appendix B

### Data extraction form

|  |  |
| --- | --- |
| Study and design general information | Design<br>Publication date<br>Journal<br>Authors and countries<br>Sample size |
| Objective 1:<br>How physiotherapy services and SDT intervention were delivered | Physiotherapy services<br>in-person,<br>telerehabilitation,<br>hybrid<br>Describe the type of delivery:<br>SDT intervention<br>in-person,<br>telerehabilitation,<br>hybrid<br>Describe the type of delivery: |
| Objective 2:<br>WHY? types of outcomes being targeted | Motivation<br>types of motivation (e.g., autonomous, extrinsic...)<br>Adherence<br>how was it measured?<br>Effectiveness<br>how was it measured?<br>health-equity<br>how was it measured? |
| Objective 3:<br>WHO, the population and types of health conditions | Population<br>Age<br>Sexe and or gender<br>Culture?<br>Other characteristics<br>Health conditions |

